# Acute nasal dryness in COVID-19

**DOI:** 10.1101/2020.11.18.20233874

**Authors:** Jordi Navarra, Alba Ruiz-Ceamanos, Juan José Moreno, José María García-Basterrechea, Josep de Haro-Licer, Scott Sinnett, Charles Spence, David Gallardo-Pujol

**Author notes:** **Corresponding author**: Jordi Navarra, Department of Cognition, Development and Educational Psychology, University of Barcelona, Spain, Faculty of Psychology, Passeig de la Vall d’Hebron, 171, 08035, Barcelona, Spain.

## Abstract

One of the entry routes of SARS-CoV-2 is the nasal epithelium. Although mounting evidence suggests the presence of olfactory dysfunction, and even anosmia, in patients with COVID-19, it is not clear whether these patients also suffer from other “nasal” symptoms that may influence their olfaction. A group of 35 patients with COVID-19 (and a control group matched in gender and age) were surveyed about the presence of a variety of nasal symptoms that may be associated to drastic perturbations experienced in the nasal cavity (e.g., “excessive dryness” and/or a continual sensation of having had a “nasal douche”). We used a cross-sectional, retrospective survey, targeted at the general population by means of non-quoted, non-random, snowball sampling. Symptoms were assessed with absence/presence responses. The possible association between two continuously distributed latent variables from categorical variables was estimated by means of polychoric correlations. More than 68% of the patients reported at least one “nasal” symptom. The clinical group also experienced “a strange sensation in the nose” and having excessive nasal dryness significantly more often than the control group. Fifty-two percent of the patients (but only 3% of the control group) reported a constant sensation of having had a strong nasal douche. Nasal symptoms predominantly co-occurred with anosmia/hyposmia, and ageusia/hypogeusia, appeared principally before or during the other symptoms of COVID-19, and lasted for twelve days, in average. The presence of these nasal symptoms, and their early occurrence, could potentially facilitate early diagnosis of COVID-19 and initial social distancing efforts.

## Introduction

In 2020, a new disease, known as COVID-19, caused by an infection of SARS-CoV-2, has spread globally. Given the high percentage of patients with COVID-19 who are asymptomatic or experience only mild symptoms (Kim et al., 2020), the need to identify (and isolate if necessary) possible carriers of the SARS-CoV-2 virus quickly and inexpensively is crucial to helping reduce its spread. Recognizing and understanding all of the possible symptomatic manifestations of COVID-19, including those that seem to be less life-threatening, can be relevant for diagnostic, treatment, and mitigation efforts (e.g., social distancing), especially in situations where RT-PCR tests cannot be administered to all non-severe cases. Along these lines, we now know, for example, that approximately 80% of COVID-19 patients in Europe report some loss of smell and taste (Lechien, Chiesa-Estomba, De Siati, 2020; see also Moein, Hashemian, Mansourafshar, Khorram-Tousi, Tabarsi, and Doty, 2020).

Evidence suggests that the virus adheres to angiotensin-converting enzyme 2 (ACE-2) receptors in epithelial cells (Wan, Shang, Graham, Baric, and Li, 2020). Clear symptomatic manifestations (e.g., loss of smell; see Brann, Tsukahara, and Weinreb, 2020; Lechien et al., 2020; Moein et al., 2020; Spinato, Fabbris, and Polesel, 2020) associated with damage in nasal (Sungnak et al., 2020; Ziegler, Allon, and Nyquist, 2020), and/or brain cells (Baig, Khaleeq, Ali, and Syeda, 2020; Mao et al., 2020) expressing ACE-2 have been identified. Goblet cells have recently been shown to harbor ACE-2 proteins (e.g., Ziegler et al., 2020). These epithelial cells are responsible for the production of the mucus in respiratory, reproductive, and intestinal tracts. Recent evidence also points to the presence of goblet secretory cells with ACE-2 membrane proteins in the respiratory nasal epithelium (e.g., Sungnak et al., 2020). A possible disruption in the mucus production by the action of SARS-CoV-2 on these cells may imply a drastic change of the mucosa environment. As sudden changes in mucus density could compromise the final adherence of the volatile chemical compounds to their corresponding odorant receptors, olfaction may possibly be compromised in the process (Hummel, Whitcroft, and Andrews, 2016).

Most important for our purpose, an ostensible reduction of mucus could certainly lead to strange sensations in the nasal cavity (Hildenbrand, Weber, Brehmer, 2011). If patients with COVID-19 do experience odd, and even unprecedented sensations in their nasal cavity, we might be observing yet another warning sign of COVID-19 that has been ignored so far, probably due to its being overshadowed by more severe symptoms needing urgent intervention.

The goal of the present study was to obtain diagnostic-valuable data regarding the presence of these non-olfactory nasal symptoms in patients with COVID-19. The study was conceived as a cross-sectional, retrospective survey targeted to the general population by means of non-quoted, non-random, snowball sampling. A brief online survey gauged whether participants had noticed any strange sensations in their nose/nasal cavity or not. More specifically, questions addressed if they felt dryness in their nasal cavity or had perceived the continuous feeling of having had a nasal douche. Information regarding the time of appearance of these symptoms relative to other COVID-19 symptoms was also gathered.

## Materials and Methods

The study was approved by the University of Barcelona’s Bioethics Commission (CBUB) and followed the Declaration of Helsinki ethical standards and the Spanish and EU data protection regulations. Written informed consent was obtained prior to participation in the survey.

### Data Collection

Information regarding the presence/absence of non-olfactory nasal symptoms, as well as regarding the presence/absence of other symptoms previously described in the literature (e.g., fever, cough, smell and taste loss, etc.), was obtained from a group of patients with COVID-19 and a control group.

Data were collected by means of an online (Qualtrics) survey, which was designed to: 1) capture the main demographics of respondents, 2) ask whether they were COVID-tested or COVID-suspected (as suspicion without a PCR test result was considered an exclusion criterion), and 3) collect information about COVID-19 symptoms.

The survey questionnaire comprised a total of 69 items in four different languages (Catalan, Spanish, Italian, and French) and were administered depending on the participants’ responses (i.e., some questions appeared only in cases where a specific response to a particular item was provided by the participant). Snowball sampling was used to recruit the sample of participants (Goodman, 1961).

### Participants

Only those participants who provided a copy of their positive result for SARS-CoV-2 in a PCR test were included in the patient group. The survey was conducted from 03/30/2020 to 04/06/2020 and a total of 414 people took part.

The vast majority of respondents were from Spain (86%), followed by Italy (9%) and France (1.5%). Responses from the survey were used to classify respondents as either patients (with demonstrable positive results in a PCR test), controls, or those with an uncertain status. Data from the latter were removed from the analyses.

The clinical group of patients with confirmed positive RT-PCR test results included 35 participants (24 female; range=21-65 years; median age=47 years; interquartile range (IQR)= 38.5-53.5 years), and the control group included 156 participants (110 female; range=16-76 years; median age= 31 years; IQR=19-50 years). Because these initial groups differed in terms of age (p<.001) and smoking habits (p<.01), only responses from a sub-sample of 35 healthy volunteers (based on the following formula: *Group∼Gender+Age+Smoking*) were used for statistical analyses, ensuring that participants were matched in terms of age (clinical group’s median age=47 years, IQR=38.5-53.5; control group’s median age=46 years, IQR=38.5-53.5), and gender (24 vs. 19 female, respectively; see Table 1 for basic demographic information). However, the groups were not completely matched in terms of smoking habits (p=.004), given that no patients (but 8 participants from the control group) reported smoking. The distribution of overall symptoms for both groups can be found in Table 2.

**Table 1.** Basic demographics and characteristics of the clinical and control groups.

|  | **Clinical**<br>**(N=35)** | SD (or %) | **Control**<br>**(N=35)** | SD (or %) |
| --- | --- | --- | --- | --- |
| Age (yrs) | 46 | 11 | 45.9 | 11 |
| Females | 24 | 68.6 | 19 | 54.3 |
| Smoking prevalence |  |  |  |  |
| No | 35 | 100 | 27 | 77.1 |
| < 10 cigarettes/day | 0 | 0 | 6 | 17.1 |
| >10 cigarettes/day | 0 | 0 | 2 | 5.8 |

**Table 2.** Distribution of symptoms in clinical and control participants.

|  | **Clinical**<br>**(N=35)** | **%** | **Control**<br>**(N=35)** | **%** |
| --- | --- | --- | --- | --- |
| Low-grade fever | 28 | 82.4 | 2 | 6.06 |
| Fever | 19 | 55.9 | 0 | 0.00 |
| Dry cough | 25 | 73.5 | 8 | 24.24 |
| Respiratory difficulty | 20 | 58.8 | 1 | 3.03 |
| Sore throat | 12 | 35.3 | 9 | 27.27 |
| Nasal congestion | 19 | 55.9 | 10 | 30.30 |
| Headache | 30 | 88.2 | 11 | 33.33 |
| Fatigue | 33 | 97.1 | 2 | 6.06 |
| Diarrhea | 20 | 58.8 | 5 | 15.15 |
| Anosmia | 29 | 85.3 | 1 | 3.03 |
| Ageusia | 27 | 79.4 | 3 | 9.09 |
| Some nasal disturbances | 19 | 55.9 | 4 | 12.12 |
| Nasal dryness | 21 | 61.8 | 5 | 15.15 |

|  | Clinical<br>(N=35) | % | Control<br>(N=35) | % |
| --- | --- | --- | --- | --- |
| Nasal douche | 17 | 51.5 | 1 | 3.03 |

### Statistical analyses

The statistical software R 4.0 (package e1071) was used to match cases and controls and to analyze the data. Values are reported as median (IQR). Comparisons of proportions were assessed using a Fisher’s exact 2-tailed test. Differences in the duration of the symptoms were tested with an unpaired 2-tailed Welch’s t test, and 95% CIs are reported. The possible co-occurrence of symptoms was calculated on the basis of correlations for categorical variables. These correlations were computed via the “polychoric” function of the psych package using frequencies as input data. The significance level was set at p < .05, and uncertainty estimates are given as confidence intervals.

## Results

The participants’ responses revealed that 68.6% of the patients experienced at least one of these “nasal” symptoms (Figure 1A). Further analyses showed that the clinical group reported having experienced excessive nasal dryness significantly more often than the control group (21 (61.8%) vs. 5 (15%); odds ratio, 8.7; 95% CI, 2.5-36.5 p<.001; see Figure 1B). Similarly, the clinical group also differed statistically from the control group in terms of having “strange nasal sensations” (19 (55.9%) vs. 4 (12%); odds ratio, 37.2, 95% CI, 5-1,666.4; p<.001; see Figure 1B). Further, while nearly half of the patients (17) reported a sensation similar to that of having had a strong nasal douche, only a single control participant reported such a sensation (1 (3%); odds ratio, 32.3; 95% CI, 4.3-32.3; p<.001; see Figure 1B).

**Figure 1.**
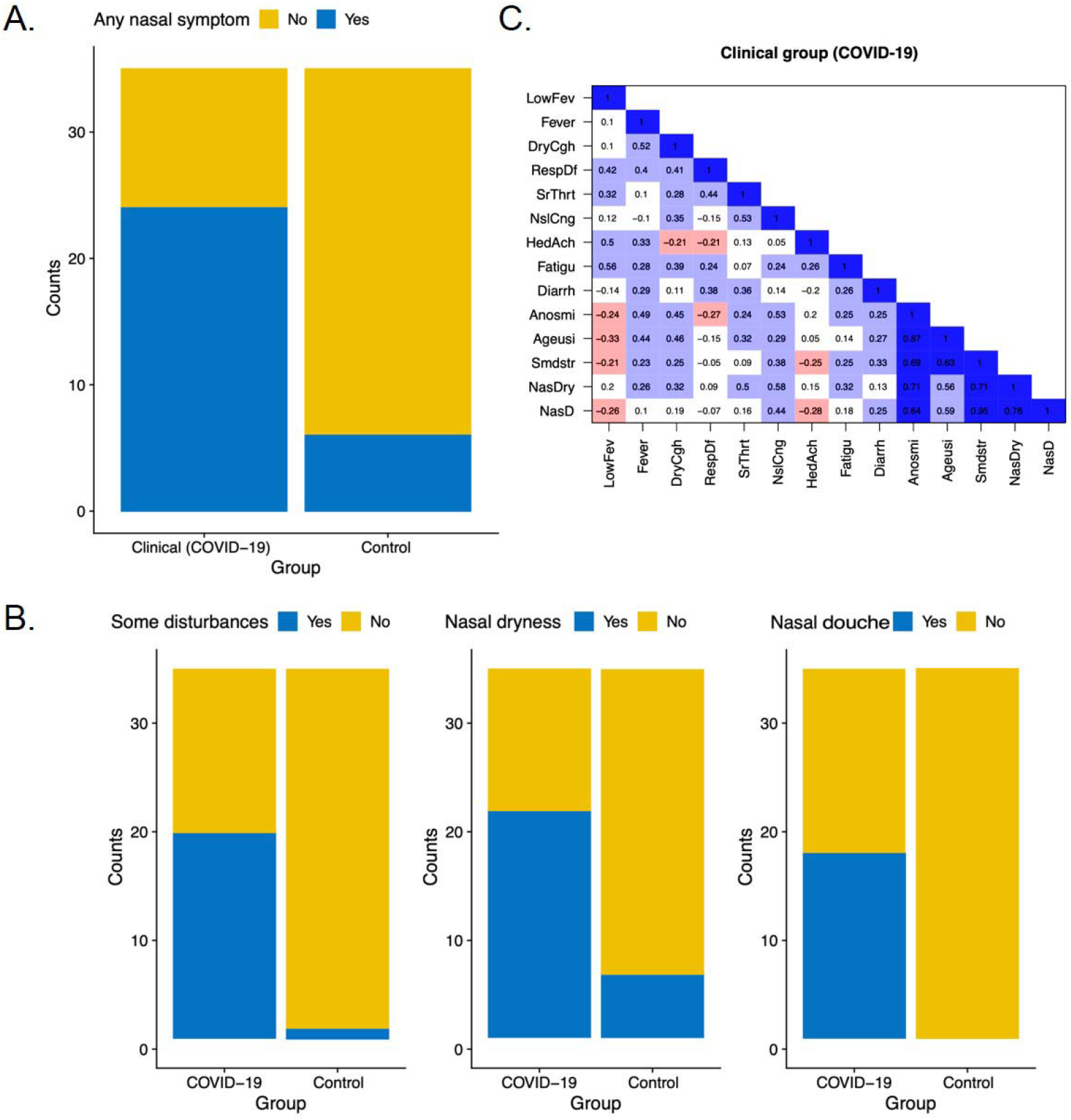
(legend). Distribution of subjective “nasal” symptoms in each group, and correlations between nasal and other symptoms. The clinical and control groups differed statistically in (A) the presence of nasal symptoms in general. (B) The 2 groups also differed in their subjective perception of three different “nasal” symptoms: Some nasal disturbances, nose dryness, and nasal douche sensation. (C) Co-occurrence matrices of symptoms for the clinical group (COVID-19). LowFev=Low grade fever; DryCgh=Dry cough; RespDf=Respiratory difficulties; SrThrt=Sore throat; NslCng=Nasal congestion; HedAch=Headache; Fatigu=Fatigue; Diarrh=Diarrhea; Anosmi=Anosmia; Ageusi=Ageusia; Smdstr=Some disturbances; NasDry=Nasal Dryness; NasFl=Nasal Flush.

The nasal symptoms reported by the patients tended to co-occur mostly with anosmia/hyposmia, and ageusia/hypogeusia (see Figure 1C), appeared before or during (but not after) other symptoms of COVID-19 (χ2= 15.55, df=3, p<.01), and lasted for about 12.05 days in patients versus 5 days in controls (p=.04), on average (95% CI of the difference=3.9-10.18). It is worth highlighting that 85.3% and 79.4% of the patients with COVID-19 reported smell and taste loss, respectively, to some extent, in line with recent literature (see Brann et al., 2020; Lechien et al., 2020; Moein et al., 2020; Spinato et al., 2020).

## Discussion

The pattern of results reveals an abnormal presence of subjective nasal symptoms in a group of patients who tested positive for COVID-19. While the presence of these sensations are likely irrelevant with respect to patient outcome (though some patients described them as extremely annoying), the early presence of these symptoms and the fact that they are clearly distinguishable from the loss of smell and taste previously reported in the literature (e.g., Spinato et al., 2020) highlight the potential importance of their use when exploring protocols for COVID-19. It is, however, plausible that all nasal and olfactory manifestations of the illness have related etiologies (i.e., impairment of epithelial cells that express ACE-2), as sudden changes in mucus density (due to the impairment of nasal goblet cells) not only induce nasal dryness (Hildenbrand et al., 2011), but also compromise the final adherence of the volatile chemical compounds to their corresponding odorant receptors (see Hummel, 2016). The presence of these nasal sensations could be taken into account for both diagnostic and social distancing purposes, especially in those situations in which RT-PCR tests cannot be administered to non-severe cases.

A possible limitation of the present study is the possibility of having false COVID-19 negative participants in the control group. It is worth mentioning that any study that tests only for patients might result in a false-negative proportion, but given the seroprevalence in the Spanish population (5%) at the time of this study, this might imply the addition of roughly 8-9 more participants with positive result in PCR test in our full sample, or 1-2 in the subsample, which would not alter the results significantly. Considering that nasal dryness could easily be related to some initial damage caused by SARS-CoV-2, future studies might determine the point in time where patients experiencing abnormal nasal dryness begin to develop other COVID-19 symptoms.

## Data Availability

The data is available in the following repository:
Open Science Framework
https://osf.io/rwpjb/

## Acknowledgments

We would like to thank all of the participants that volunteered to answer the survey used to obtain the data reported in the present study.

